# Knowledge of COVID-19 Symptoms, Transmission, and Prevention: Evidence from Health and Demographic Surveillance in Southern Mozambique

**DOI:** 10.1101/2023.03.31.23288026

**Authors:** Ariel Q. Nhacolo, Zachary J. Madewell, Jonathan A. Muir, Charfudin N. Sacoor, Elisio G. Xerinda, Teodimiro Matsena, Quique Bassat, Cynthia G. Whitney, Inácio M. Mandomando, Solveig A. Cunningham

## Abstract

Over 230,000 COVID-19 cases and over 2,200 deaths have been reported in Mozambique though May 2023. Understanding community members’ knowledge of SARS-CoV-2 transmission and prevention is essential for directing public health interventions to reduce disease spread and improve vaccination coverage. Here, we describe knowledge of COVID-19 transmission, prevention, and symptoms among community residents in Mozambique. We conducted a cross-sectional survey among 33,087 households in a Health and Demographic Surveillance System in Manhiça, Mozambique. Participants were recruited at the tail end of the Delta variant wave in September 2021 to the peak of Omicron cases in January 2022. Principal components analysis was used to create scores representing knowledge of COVID-19 symptoms, transmission, and prevention. Multiple imputation and quasi-Poisson regression were used to examine associations between demographic characteristics and sources of COVID-19 information, and knowledge of COVID-19 symptoms, transmission, and prevention. We examined whether sources of COVID-19 information mediated the relationship between educational attainment and knowledge of symptoms, transmission, and prevention. Across this rural community, 98.2%, 97.0%, and 85.1% of respondents reported knowing how COVID-19 could be prevented, that SARS-CoV-2 can cause disease, and how SARS-CoV-2 is transmitted, respectively. Most cited symptoms were cough (51.2%), headaches (44.9%), and fever (44.5%); transmission mechanisms were droplets (50.5%) or aerosol (<5 µm diameter) (46.9%) from an infected person; and prevention measures were handwashing (91.9%) and mask-wearing (91.8%). Characteristics associated with greater knowledge of symptoms, transmission, and prevention included having at least primary education, older age, employment, higher wealth, and Christian religion. Respondents who had had COVID-19 symptoms were also more likely to have knowledge of symptoms, transmission, and prevention. Gathering information from TV, WhatsApp, radio, and hospital mediated the relationship between educational attainment and knowledge scores. These findings support the need for outreach and for community-engaged messaging to promote prevention measures, particularly among people with low education.

## Introduction

Coronavirus disease 2019 (COVID-19) was first reported in Mozambique in March 2020. Since then, there have been over 230,000 confirmed cases and over 2,200 deaths reported in Mozambique through May 2023 [1], though the death toll could be much higher than official figures due to the challenges experienced in the country’s surveillance capacities and under-reporting [2].

Effective public health education programs are required to reduce the burden of COVID-19 and the strain on healthcare system resources. Studies of COVID-19 in sub-Saharan Africa have demonstrated positive associations between higher level of knowledge and practicing prevention measures [3, 4]. Knowledge of COVID-19 transmission and disease was also positively associated with vaccination coverage [5], which is one of the most effective strategies for protecting individuals against COVID-19 hospitalization and death [6, 7]. As of May 2023, 59.1% of Mozambique’s population had completed a primary COVID-19 vaccine series of BBIBP-CorV (Sinopharm, Beijing CNBG) (two doses), Ad26.COV2.S (Janssen) (one dose), or ChAdOx1-S (Covishield) (two doses), but only 3.5% of these had received a booster dose [1]. COVID-19 vaccines became available for adolescents aged 12-17 in Mozambique in November 2022 [8].

Currently, there is limited information about knowledge of COVID-19 transmission, prevention, and symptoms in Mozambique. Two cross-sectional surveys on knowledge, attitudes and practices (KAP) among community healthcare workers in Mozambique using an online health platform [9] and among adolescents using a cross-sectional survey in two provinces of Central Mozambique [10] found low knowledge of symptoms, transmission, and prevention measures, but these were done early in the pandemic, before the emergence of more transmissible variants such as Delta and Omicron [11, 12]. The studies’ generalizability was also limited by convenience samples and small sample sizes. We conducted a population-based, representative study to evaluate factors associated with knowledge of COVID-19 symptoms, transmission, and prevention, in a rural setting of southern Mozambique. This approach is important for reducing household transmission and improving vaccination coverage [13] and provides information on how to tailor communication strategies to reduce community infectious disease transmission.

## Materials and Methods

### Study design

This study is part of a broader examination within the Child Health and Mortality Prevention Surveillance (CHAMPS) network to analyze the consequences of COVID-19 lockdowns for child health and mortality [14–17]. Leveraging the established platform of the CHAMPS Network of Health and Demographic Surveillance Systems (HDSSs) [18], we administered a short questionnaire to all households in a rural district [19]. The Strengthening the Reporting of Observational Studies in Epidemiology (STROBE) reporting guidelines for cross-sectional studies were followed.

### Study setting and population

Manhiça is a district in Maputo Province, located about 85 km north of the capital city, Maputo. An HDSS was established there in 1996 by the Manhiça Health Research Center (CISM) and currently covers the entire Manhic_Ja District, which spans 2,380 km^2^. More information about the Manhic_Ja HDSS has been published elsewhere [20]. With a population of 201,845 [20], Manhiça is the second most populated district in Maputo Province after the Matola District (the capital of Maputo Province). The major economic activity is sugarcane farming—Maragra and Xinavane are the main sugar factories in the country. The health system in Manhiça comprises one district hospital located in Manhiça village, one rural hospital located in the Xinavane administrative post, and 19 health centers.

Manhiça is a geographic corridor highly exposed to transmissible diseases such as HIV, and currently COVID-19, stemming from population migration. A large proportion of the Manhiça population migrate to the nearby capital city of Maputo or beyond to South Africa with regular return visits to their households in Manhiça. Moreover, the district is crossed by Mozambique’s main road (National Road Number 1), which connects the Southern, Central, and Northern regions of the country as well as neighboring countries of South Africa, Eswatini, Zimbabwe, Malawi, Zambia, and Tanzania. The district is also crossed by a railroad that connects to Maputo city and harbor. These transportation systems facilitate population migration and disease spread.

In each household, one household member was asked to participate on behalf of the household. Household members were eligible for inclusion if they resided at the residence between March 2020 and the date of interview between September 2021 and January 2022. The questionnaire was conducted together with standard procedures for regular HDSS visits to all residences in the defined catchment area (census method) [20].

### Data collection and quality assurance

A survey instrument was developed to collect information about households’ experiences during the COVID-19 lockdown, which included information on knowledge regarding COVID-19 symptoms, transmission, and prevention. Questions were based on guidelines issued by the Ministry of Health for COVID-19 prevention in Mozambique [21]. Interviewers asked open-ended questions and recorded participants’ responses. Data collection was conducted by HDSS fieldworkers during their regular visits to the households between September 2021 and January 2022 through tablet-based in-person interviews with heads of households or their representatives if the heads were unavailable. These data were linked with data from the HDSS questionnaires to incorporate socio-demographic data about the households: household size, number of children under 5 years of age, number of adults over 60 years, and number of pregnant women; household assets; and materials used for constructing the houses (Supplemental Methods, S1-S2 Figs); variables about the head of household were age, sex, occupation, education, religion, mother language, and marital status.

Fieldworkers and supervisors were trained on the use of this new module by the study coordinator. Data cleaning and quality assurance followed standard procedures for the HDSS, whereby 5% of households visited each week were revisited to confirm the recorded information [22]. There was a script to filter errors at the time of uploading the data from the tablets to the server. Records with inconsistences were returned to the field for reconciliations.

### Measures

Head of household variables were: sex (female, male), age group (<18 years, 18-39 years, 40-64 years, ≥65 years), religion (Catholic, Protestant, Christian unspecified, Zion church member, atheist, Evangelical, other, don’t know), language (Tsonga, Echuwabo, Cisena, Bitonga, other), education (higher education, technical education, secondary education, primary education, no education), occupation (retired/pensioner, does not work, professional, merchants, skilled manual, unskilled manual, student/volunteer, other), marital status (single, married/de facto union, separated/divorced, widowed), and had had COVID-19 symptoms since COVID-19 was first reported in Mozambique (yes, no). Household variables were: household size (1, 2, 3, 4, 5, 6+), number of children under 5 years (0, 1, 2+), number of adults over 60 years (0, 1, 2+), number of pregnant women (0, 1+), and wealth index, which was derived from principal components analysis (PCA) (Supplemental Methods).

In accord with other KAP studies [17, 23–25], we used PCA-based factor analysis to create scores for assessing the degree of knowledge of COVID-19 symptoms, transmission, and prevention (Supplemental Methods). The overall Kaiser-Meyer-Olkin index of sampling adequacy was 0.71, 0.73, and 0.83 for knowledge of symptoms, transmission, and prevention, so we concluded the sample size and data were adequate for the PCAs. The resultant compound factor for knowledge of symptoms included seven variables that accounted for 32% of the variability in the data: difficulty breathing, dry cough, fever, headaches, muscle pain, and sore throat (Table S1). The compound factor for knowledge of transmission included eight variables that explained 29% of the variation: hugging, kissing, and droplets from an infected person; and touching a fomite (objects or materials which are likely to carry infection, such as clothes, utensils, and furniture), an infected person, or one’s own eyes or nose, or mouth. The compound factor for knowledge of prevention included eight variables that explained 31% of the data variability: avoid crowded places, touching eyes, touching mouth, touching nose, or traveling; social distancing; quarantine; and wash hands with alcohol. Knowledge of symptoms, transmission, and prevention scores ranged from 0 to 3.3, 0 to 4.2, and 0 to 4.7, respectively, with higher scores representing greater knowledge.

Our survey also asked participants what they would do if someone in their family had symptoms suggestive of COVID-19. Response categories included: go to the hospital, quarantine, call the hospital, call the community leader, and treat symptoms at home; respondents could have said yes to multiple categories. Sources of information about COVID-19 included: TV, SMS/WhatsApp, radio, hospital, community leaders.

### Statistical Analysis

We reported frequency distributions of individual characteristics (age, sex, education, occupation, religion, language, marital status) and household characteristics (wealth index, total number of household members, number under 5 children, number above 60 adults, number of pregnant women). We also reported frequencies and 95% confidence intervals (95% CI) for knowledge of COVID-19 symptoms, transmission, and prevention; management of suspected cases; and sources of information. There were minimal missing data for demographic variables, between 0.1% (age) to 9.6% (education). Consistent with other KAP studies [26–29], we used multiple imputation with predictive mean matching for these missing data to retain statistical power and avoid selection bias (Supplemental Methods).

Quasi-Poisson regression was used to evaluate unadjusted and adjusted associations between characteristics (age; sex; language; religion; marital status; education; occupation; wealth index; had COVID-19 symptoms; number of household members, children under 5, older adults, and pregnant women; sources of information) and knowledge of symptoms, transmission, and prevention scores (Supplemental Methods). Logistic regression was used to evaluate unadjusted and adjusted associations between the same characteristics and whether the respondents had experienced symptoms associated with COVID-19. Finally, we assessed whether sources of COVID-19 information mediated the relationship between educational attainment and knowledge of symptoms, transmission, and prevention scores (Supplemental Methods). This analysis followed causal mediation analysis methods as previously described by VanderWeele [30] and has been used in other KAP studies [31–34]. All analyses were done in R software, version 4.2.3 (R Foundation for Statistical Computing, Vienna, Austria).

### Ethical statement

The HDSS data collection has ethical approval from the Institutional Ethics Review Board for Health (CIBS) and Internal Scientific Committee (CCI) at CISM, and the National Bioethics Committee for Health (CNBS-Mozambique).

This study used part of the existing HDSS data, for which all the heads of households and household members in Manhiça district have voluntarily agreed and signed a written detailed informed consent for providing their demographic and socio-economic data, including that of their households and their young dependents (children under the age of 18 years), in the context of HDSS. In relation to new data, the study team obtained approval from CISM’s CCI and CIBS to collect the data based on voice-recorded informed oral consent to minimize the risk of COVID-19 transmission when handling paper-based informed and signed consents between interviewers and interviewees. Data collection was mainly over the phone to minimize contacts due to COVID-19. Even during household visits (which were done for participants unreachable by phone), informed consent was oral and voice-recorded to minimize the risk of COVID-19. Written informed consent was obtained from the parent or guardian of each participant under 18 years of age.

All communications with study participants were done with the language of each participant’s preference. Where the preferred language was not Portuguese, the fieldworkers translated the questionnaires *in situs* as in other studies that the HDSS and social science team has conducted. Where the fieldworker could not speak the participant’s preferred language, a translator was sought in the household, or a suitable fieldworker conducted the communication at another time. Data collection was done between September 2021 and January 2022.

## Results

The Manhiça HDSS is home to 40,636 active households in 2021, of which 33,087 (81.4%) responded to this survey (Fig 1). Respondents (18,823, 56.9%) were heads of household, their spouses (7,905, 23.9%), children (3,025, 9.1%), or other family members (3,334, 10.1%). The majority of respondents (72.4%) were female, and the median age was 38 years (interquartile range [IQR]: 27–53 years) (Table 1). More than half (54.4%) spoke Xirhonga, 34.5% were Zion church members, 70.0% had primary or no education, and 73.5% were manual laborers. Median household size was 3 (IQR: 2–5). Of all households, 43.5% had children under 5 years, 26.5% had adults over 60 years, and 3.0% had pregnant women (Table 1). Eight percent (2,465/33,087) of respondents reported having had symptoms suggestive of COVID-19 since the disease was first reported in Mozambique, most commonly flu-like symptoms (50.4%, 1,242/2,465), dry cough (48.0%, 1,182/2,465), headaches (33.8%, 832/2,465), fever (27.0%, 665/2,465), and cough with sputum (21.9%, 541/2,465) (Table S2).

**Fig 1.**
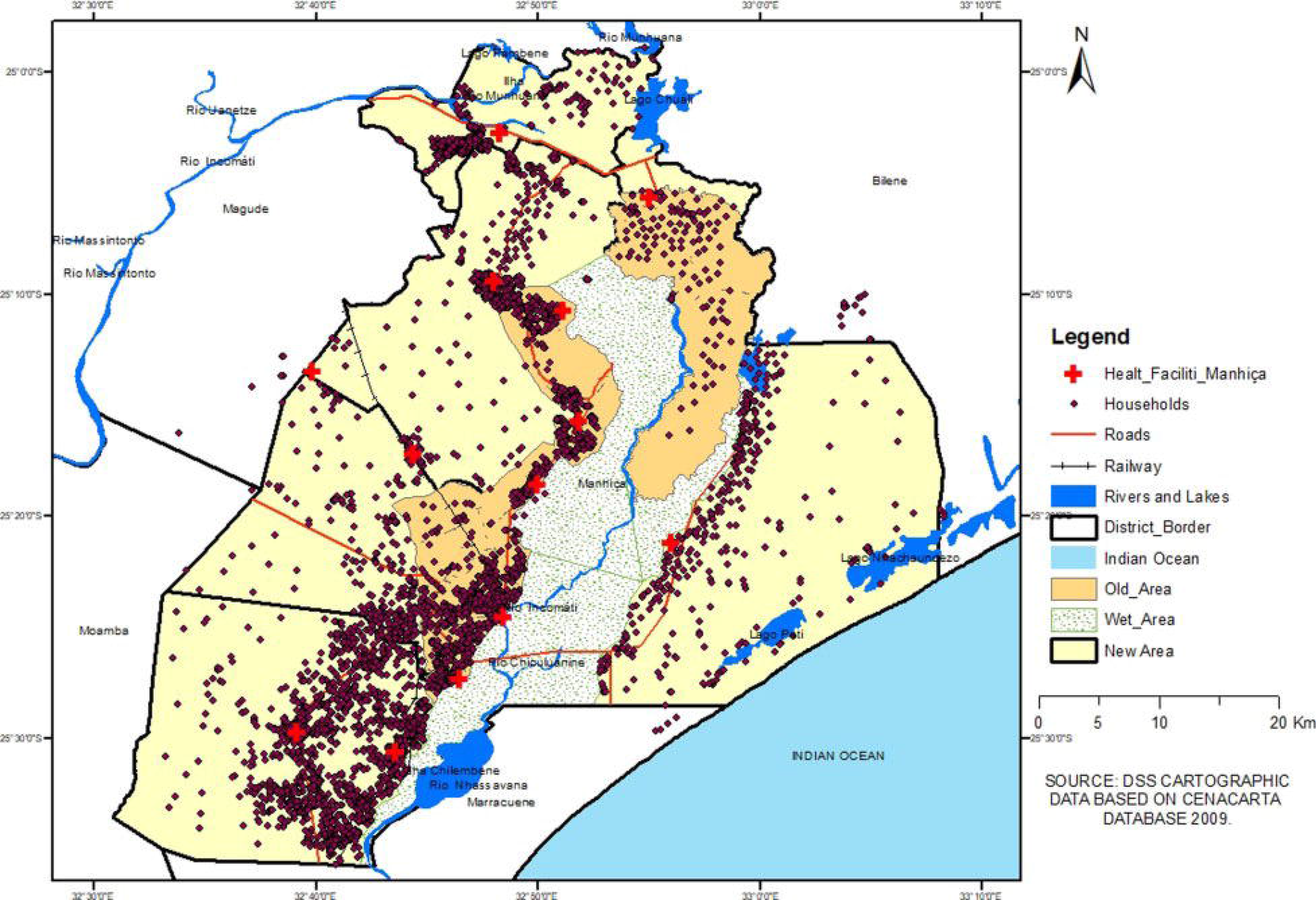
Map of Manhiça District. Source: Created by the authors using Health and Demographic Surveillance System (HDSS) cartographic data.

**Table 1.**
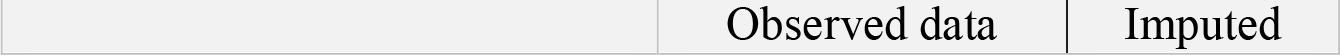
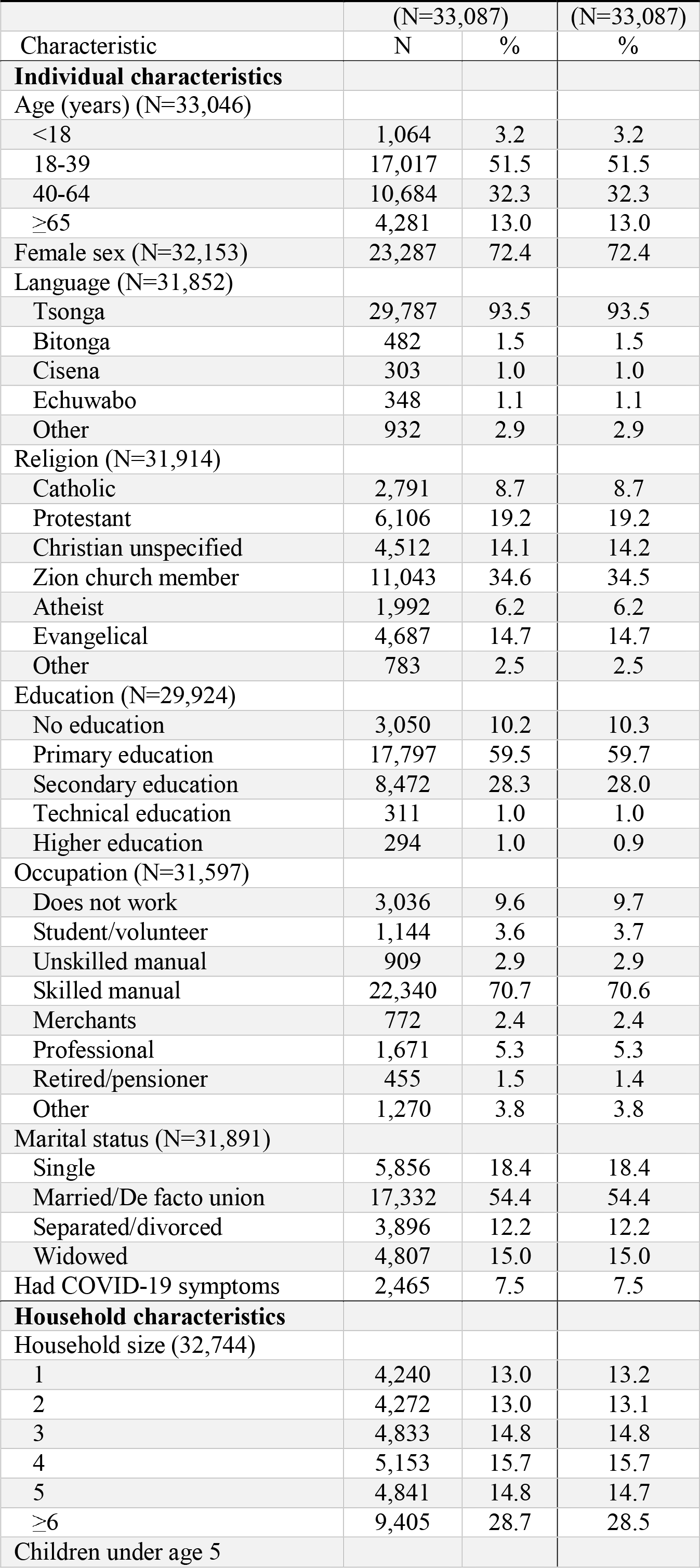
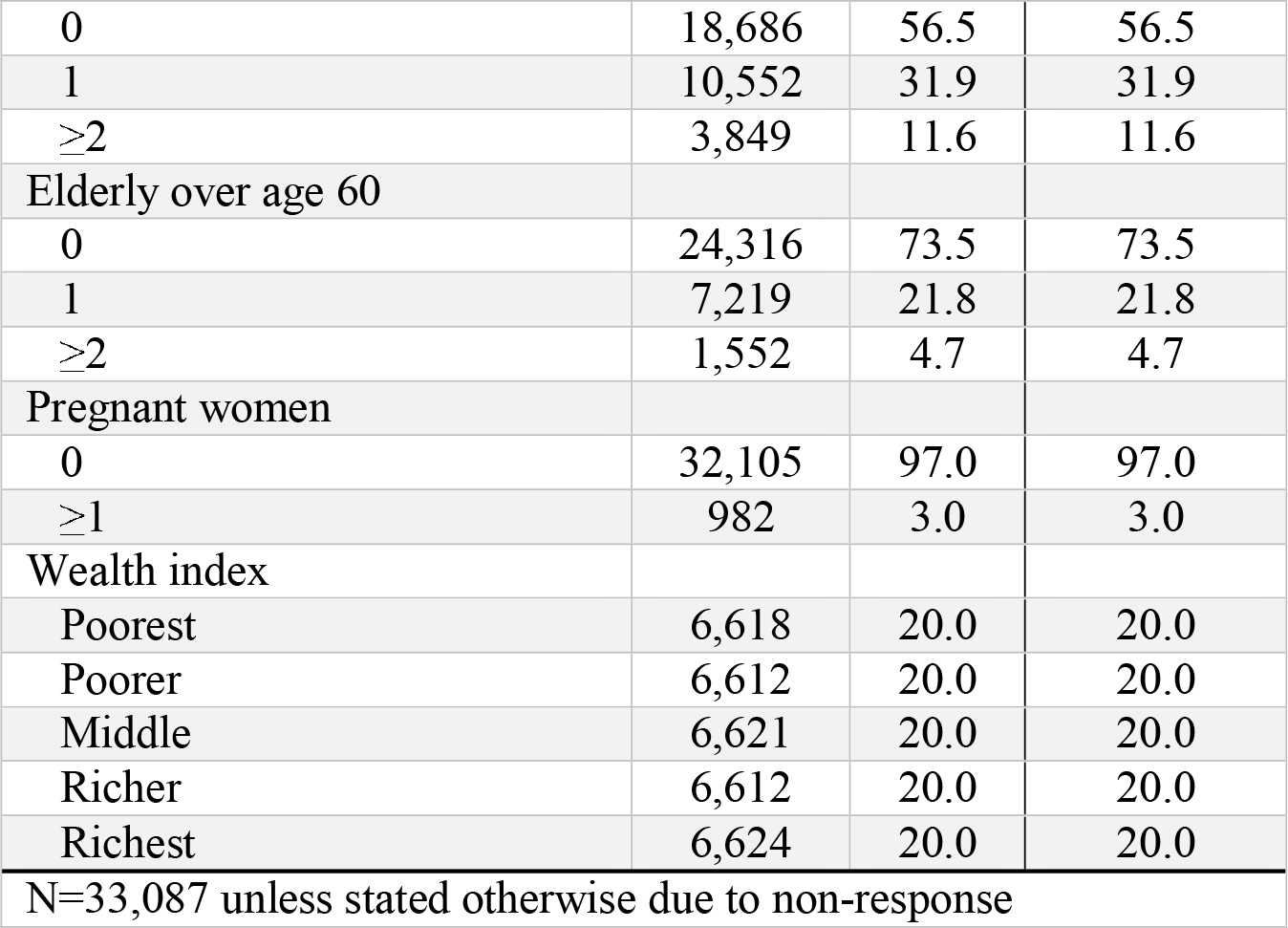
Descriptive statistics of individuals who participated in COVID-19 cross-sectional survey, Mozambique, September 2021 – January 2022, (N=33,087)

|  | (N=33,087) |  | (N=33,087) |
| --- | --- | --- | --- |
| Characteristic | N | % | % |
| **Individual characteristics** |  |  |  |
| Age (years) (N=33,046) |  |  |  |
| <18 | 1,064 | 3.2 | 3.2 |
| 18-39 | 17,017 | 51.5 | 51.5 |
| 40-64 | 10,684 | 32.3 | 32.3 |
| ≥65 | 4,281 | 13.0 | 13.0 |
| Female sex (N=32,153) | 23,287 | 72.4 | 72.4 |
| Language (N=31,852) |  |  |  |
| Tsonga | 29,787 | 93.5 | 93.5 |
| Bitonga | 482 | 1.5 | 1.5 |
| Cisena | 303 | 1.0 | 1.0 |
| Echuwabo | 348 | 1.1 | 1.1 |
| Other | 932 | 2.9 | 2.9 |
| Religion (N=31,914) |  |  |  |
| Catholic | 2,791 | 8.7 | 8.7 |
| Protestant | 6,106 | 19.2 | 19.2 |
| Christian unspecified | 4,512 | 14.1 | 14.2 |
| Zion church member | 11,043 | 34.6 | 34.5 |
| Atheist | 1,992 | 6.2 | 6.2 |
| Evangelical | 4,687 | 14.7 | 14.7 |
| Other | 783 | 2.5 | 2.5 |
| Education (N=29,924) |  |  |  |
| No education | 3,050 | 10.2 | 10.3 |
| Primary education | 17,797 | 59.5 | 59.7 |
| Secondary education | 8,472 | 28.3 | 28.0 |
| Technical education | 311 | 1.0 | 1.0 |
| Higher education | 294 | 1.0 | 0.9 |
| Occupation (N=31,597) |  |  |  |
| Does not work | 3,036 | 9.6 | 9.7 |
| Student/volunteer | 1,144 | 3.6 | 3.7 |
| Unskilled manual | 909 | 2.9 | 2.9 |
| Skilled manual | 22,340 | 70.7 | 70.6 |
| Merchants | 772 | 2.4 | 2.4 |
| Professional | 1,671 | 5.3 | 5.3 |
| Retired/pensioner | 455 | 1.5 | 1.4 |
| Other | 1,270 | 3.8 | 3.8 |
| Marital status (N=31,891) |  |  |  |
| Single | 5,856 | 18.4 | 18.4 |
| Married/De facto union | 17,332 | 54.4 | 54.4 |
| Separated/divorced | 3,896 | 12.2 | 12.2 |
| Widowed | 4,807 | 15.0 | 15.0 |
| Had COVID-19 symptoms | 2,465 | 7.5 | 7.5 |
| **Household characteristics** |  |  |  |
| Household size (32,744) |  |  |  |
| 1 | 4,240 | 13.0 | 13.2 |
| 2 | 4,272 | 13.0 | 13.1 |
| 3 | 4,833 | 14.8 | 14.8 |
| 4 | 5,153 | 15.7 | 15.7 |
| 5 | 4,841 | 14.8 | 14.7 |
| ≥6 | 9,405 | 28.7 | 28.5 |
| Children under age 5 |  |  |  |
| 0 | 18,686 | 56.5 | 56.5 |
| 1 | 10,552 | 31.9 | 31.9 |
| ≥2 | 3,849 | 11.6 | 11.6 |
| Elderly over age 60 |  |  |  |
| 0 | 24,316 | 73.5 | 73.5 |
| 1 | 7,219 | 21.8 | 21.8 |
| ≥2 | 1,552 | 4.7 | 4.7 |
| Pregnant women |  |  |  |
| 0 | 32,105 | 97.0 | 97.0 |
| ≥1 | 982 | 3.0 | 3.0 |
| Wealth index |  |  |  |
| Poorest | 6,618 | 20.0 | 20.0 |
| Poorer | 6,612 | 20.0 | 20.0 |
| Middle | 6,621 | 20.0 | 20.0 |
| Richer | 6,612 | 20.0 | 20.0 |
| Richest | 6,624 | 20.0 | 20.0 |
| N=33,087 unless stated otherwise due to non-response |  |  |  |

Nearly all (99.4%, 32,901/33,087) participants had heard of coronavirus, but 20.9% (6,926/33,087) were unfamiliar with the term COVID-19. Of all respondents, 98.2% reported knowing how SARS-CoV-2 transmission could be prevented, 97.0% knew SARS-CoV-2 may cause disease, and 85.1% knew how SARS-CoV-2 is transmitted. The most commonly mentioned prevention measures were washing hands with soap (91.9%) and wearing a facemask (91.8%), whereas least mentioned included avoiding touching eyes (3.8%), nose (4.0%), and mouth (4.9%) (Fig 2). Most mentioned transmission mechanisms were droplets (50.5%) and aerosol (<5 µm diameter) (46.9%) from an infected person; least mentioned were touching eyes or nose (9.2%), or mouth (10.7%). The most recognized COVID-19 symptoms were dry cough (51.2%), headaches (44.9%), and fever (44.5%); least mentioned were nausea/vomiting (3.7%) and muscle or body aches (13.7%). Most participants (88.6%) indicated they would take symptomatic family members to the hospital for treatment, whereas 3.8% stated they would treat symptoms at home. The most cited sources of information regarding COVID-19 were television (44.0%), community leaders (36.2%), and radio (33.7%) (Table S3).

**Fig 2.**
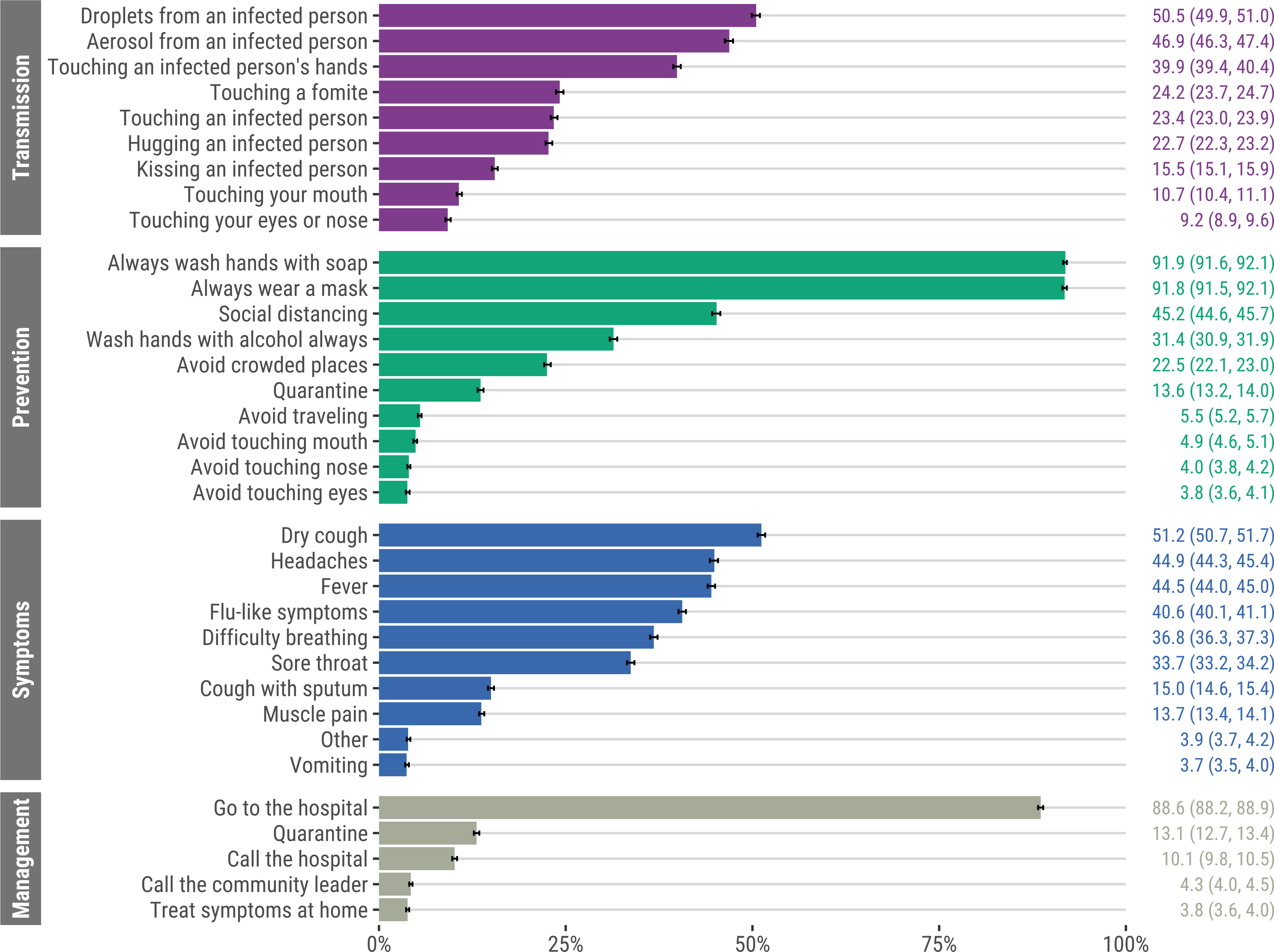
Knowledge of COVID-19 transmission, prevention and symptoms, and management of suspected cases, Mozambique, September 2021 – January 2022 (N=33,087). Error bars represent 95% confidence intervals.

Unadjusted analyses between demographics and sources of information, and PCA-derived knowledge of symptoms, transmission, and prevention scores are shown in S3 Fig: these were positively correlated (*r* = 0.47 ∼ 0.63; *p* < 0.001) (S4 Fig). Adjusting for all other variables in the models (see Fig 3), knowledge of COVID-19 symptoms, transmission, and prevention scores were highest among heads of household with higher (β coefficients: 0.43 ∼ 0.47), technical (0.42 ∼ 0.46), or secondary (0.35 ∼ 0.41) education with no education as reference; and cited TV (0.25 ∼ 0.47), radio (0.16 ∼ 0.38), or SMS/WhatsApp (0.16 ∼ 0.36) as sources of information for COVID-19. Sources of information were significant mediators of the relationship between educational attainment and knowledge of symptoms, transmission, and prevention scores (Tables S4-S6). Compared to participants with no formal education, those with ≥primary education were more likely to have cited TV, hospital, radio, SMS/WhatsApp, and community leaders as sources of COVID-19 information than not cite them as sources of information; the higher the education level, the more likely participants cited these as sources of information. A significant proportion of the positive association between educational attainment and knowledge of symptoms, transmission, and prevention scores can be explained by mediators (TV, SMS/WhatsApp, and radio sources of information): the higher the education level, the greater the impact of these mediators on knowledge scores. Conversely, there was a significant negative natural indirect effect for community leaders and hospital, which indicates these sources of information may have attenuated the positive association of education on knowledge scores. Specifically, these sources of information were associated with slightly reduced COVID-19 knowledge among those with higher education.

**Fig 3.**
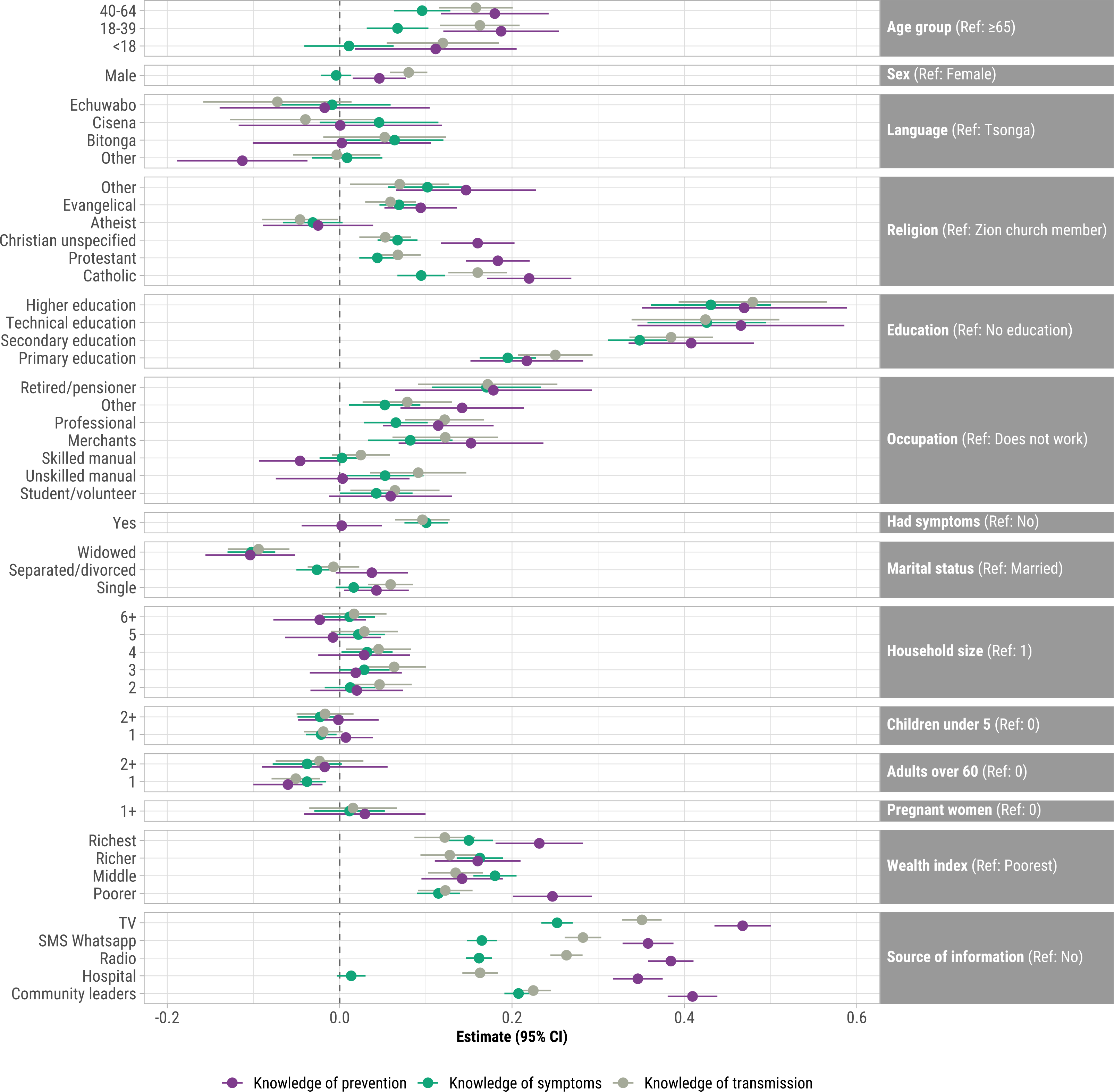
Adjusted^a^ associations between demographics and sources of information, and knowledge of prevention, symptoms, and transmission indices^b^ derived from principal components analysis, Mozambique, September 2021 – January 2022 (N=33,087). Points represent β coefficients and error bars represent 95% confidence intervals. ^a^Adjusted for all other variables in the model. ^b^Knowledge of prevention includes: avoid crowded places, touching eyes, touching mouth, touching nose, or traveling; social distancing; quarantine; and wash hands with alcohol. Knowledge of symptoms includes: difficulty breathing, dry cough, fever, headaches, muscle pain, and sore throat. Knowledge of transmission includes: avoid crowded places, touching eyes, touching mouth, touching nose, or traveling; social distancing; quarantine; and wash hands with alcohol.

In adjusted analyses, higher knowledge of symptoms, transmission, and prevention scores were among participants who were 18-64 years (0.07 ∼ 0.19) with ≥65 as reference; were Roman Catholic, Protestant, or Evangelical (0.05 ∼ 0.22) with Zion church members as reference; were merchants (0.08 ∼ 0.15), retired/pensioners (0.17 ∼ 0.18), or professionals (0.07 ∼ 0.12) with unemployed as reference; and were in the poorer to richest wealth index quintiles (0.11 ∼ 0.25) with poorest as reference (Fig 3). Knowledge scores for all three indices were significantly lower for widowed (-0.10 ∼ -0.09) compared to married participants. Furthermore, knowledge of symptoms and transmission, but not prevention, were higher for those who had COVID-19 symptoms (0.10 ∼ 0.11). Knowledge of prevention and transmission were higher for males than females (0.04 ∼ 0.08). Variance inflation factors for independent variables in all three adjusted models were <1.3, so there was no evidence of collinearity (Table S7).

The adjusted odds of having had symptoms consistent with COVID-19 were higher for participants with primary, secondary, technical, or higher education (aORs: 1.74 ∼ 6.56) with no education as reference; Catholics, Protestants, Christians, Evangelicals, and Atheists with Zion church members as reference (aORs: 1.44 ∼ 2.02); professionals (aOR: 1.30; 95% CI: 1.07-1.59) with unemployed as reference; single (aOR: 1.20; 95% CI: 1.07-1.35) with married as reference; and reported TV (aOR: 1.32; 95% CI: 1.19-1.47) as a source of COVID-19 information (Fig 4). The adjusted odds of COVID-19 symptoms were lower for individuals in the richest wealth index quintile (aOR: 0.83, 95% CI: 0.70, 0.98), were <18 years with ≥65 years as reference (aOR: 0.36, 95% CI: 0.24, 0.54) and who learned about COVID-19 from hospital, radio, or community leaders (aORs: 0.59 ∼ 0.80).

**Fig 4.**
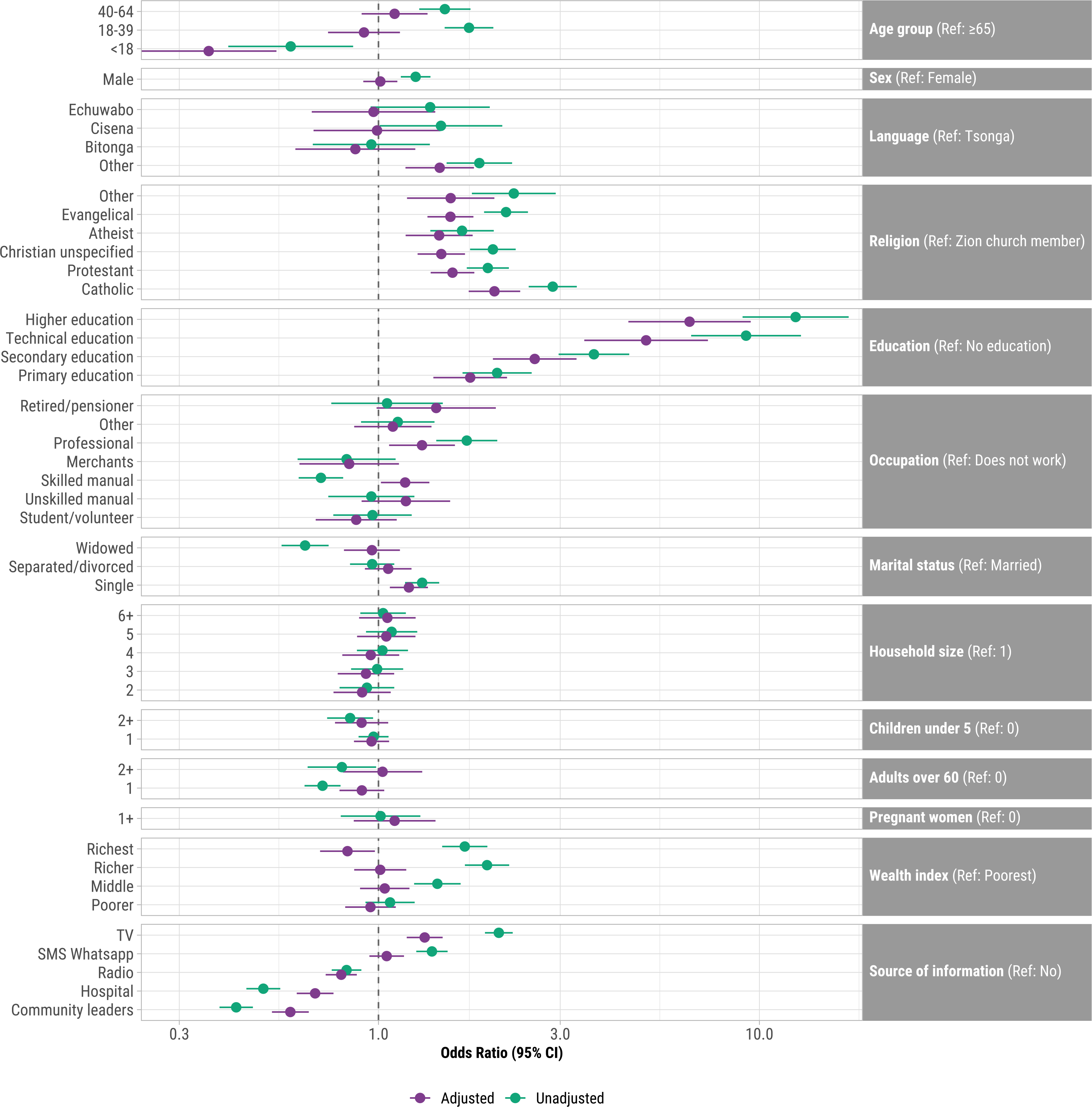
Unadjusted and adjusted^a^ associations between demographics and sources of information, and had COVID-19 symptoms, Mozambique, September 2021 – January 2022 (N=33,087). X-axis is shown on a log-scale. ^a^Adjusted for all other variables in the model.

## Discussion

Community public health measures to reduce infectious disease transmission are contingent upon individual perceptions of risk and knowledge. In this census of over 33,000 household members in Manhiça, Mozambique, almost everyone had heard of coronavirus, were aware of COVID-19 symptoms, and knew how COVID-19 could be prevented. The most recognized COVID-19 symptoms were dry cough, headaches, fever, difficulty breathing, and sore throat, consistent with findings from a systematic review of COVID-19 knowledge in sub-Saharan Africa [4]. This study was conducted at the tail end of the Delta variant wave in September through the peak of Omicron cases in January in Mozambique [35]. The most common Delta symptoms reported in the ZOE COVID Study in the U.K. included runny nose, headache, sneezing, sore throat, and loss of smell, whereas the most common Omicron symptoms were runny nose, headache, sore throat, sneezing, and persistent cough [36]. There was high knowledge of nonpharmaceutical prevention measures such as handwashing and mask-wearing, similar to a study in Ethiopia [37], but less than half of household members reported social distancing and avoiding crowded places as prevention measures, which are among the most effective public health measures to prevent SARS-CoV-2 transmission [38, 39]. Approximately half of household members knew that SARS-CoV-2 may be transmitted from droplets or aerosol from an infected person, which is the primary mode of SARS-CoV-2 transmission [40]. We also found that knowledge of symptoms, transmission, and prevention scores were positively correlated and there were consistent results between the knowledge outcomes.

The finding that participants with higher education had greater knowledge of COVID-19 symptoms, transmission, and prevention is consistent with studies in Australia [41], Ethiopia [17, 42], Indonesia [43], and South Korea [44]. Other studies also found that higher education was associated with COVID-19 vaccine acceptance [5, 45]. Mediation analyses demonstrated that TV, SMS/WhatsApp, and radio sources of information were significant mediators in the relationship between educational attainment and knowledge scores. Household heads with higher education were more likely to report TV, SMS and radio as sources of COVID-19 information, which were associated with higher knowledge scores. Only 20% of those with no education had a TV and 70% had a cellphone, whereas 86% with higher/university education had a TV and 99% had a cellphone. These resource discrepancies may also contribute to the finding that individuals in the higher wealth quintiles had higher knowledge scores. Other studies found that lower education was associated with misinformation, which may in part explain lower knowledge among this group as well [41, 46]. Participants with higher education also had the highest odds of having COVID-19 symptoms compared to those with no education, perhaps suggesting greater awareness of COVID-19 symptoms.

Older respondents had higher COVID-19 knowledge, consistent with other studies [43]. Older adults, particularly those with comorbidities, are at greater risk for severe illness, hospitalizations, and death from COVID-19: 70% of COVID-19-attributed deaths in the U.S. were among adults 70 years or older [47]. Other studies reported greater vaccination [5] and other preventive measures [48] among older adults compared to young adults. An online survey in Mozambique found that older participants were more likely to accept COVID-19 vaccines [49]. Mozambique has a young population with a median age of 17.6 years [50]. Although younger adults are less likely to be hospitalized with COVID-19 compared to older adults, some develop severe disease, and they may be infectious without symptoms [51].

Other factors associated with higher COVID-19 knowledge scores included employment as merchants, professionals, and unskilled manual laborers; and Catholic, Protestant, and Evangelical religions. Additional outreach promoting information regarding COVID-19 transmission and prevention, as well as vaccine safety and effectiveness, should be tailored to local communities in their language and should engage community leaders [29, 52]. Although not evaluated in this study, other KAP studies found higher knowledge scores among individuals with positive HIV status, urban residence, and no previous SARS-CoV-2 infection [10, 24]. Although we did not know HIV status of participants, this study was conducted in a community with high HIV prevalence. It is conceivable that previous experiences with and exposure to messaging for HIV and other endemic infectious diseases increased COVID-19 awareness in Manhiça. Additionally, individuals with HIV may have greater contact with the healthcare system, increasing the opportunity to learn about COVID-19 from healthcare providers. Rural southern Mozambique is also characterized by high levels of labor migration within Mozambique and to South Africa, but this study did not assess how that affected COVID-19 knowledge.

This study had several limitations. This study is not designed to be representative of all households in Mozambique; generalizability to a specific population is a feature of all population-based sub-national studies [18, 53]. Still, these results have broader relevance to educating communities about COVID-19 prevention. This was a cross-sectional study, which precludes establishing causal and temporal relationships between demographics and knowledge scores. There may have been social desirability bias in responses about knowledge. There may have been response bias if individuals with greater knowledge of COVID-19 were more likely to participate than those with less knowledge. Finally, there may have been recall bias due to the length of time in the study. Notwithstanding these limitations, regional census entails that these findings are representative of the district. To our knowledge, there are no other studies of knowledge of COVID-19 symptoms, transmission, and prevention among community members in Mozambique.

## Conclusions

In this census of over 33,000 community members in a rural district of Mozambique, most individuals had high knowledge of COVID-19 symptoms and prevention, but there was less knowledge about transmission. Messaging regarding COVID-19 in southern Mozambique effectively increased awareness of symptoms and prevention. These findings support the need for outreach and community engagement considering the target audience to promote COVID-19 prevention measures, particularly among vulnerable populations with lower educational status.

## Data Availability

All data produced in the present study are available upon reasonable request to the authors

## Acknowledgements

CISM is supported by the Government of Mozambique and the Spanish Agency for International Development (AECID). ISGlobal receives support from the Spanish Ministry of Science and Innovation through the “Centro de Excelencia Severo Ochoa 2019-2023” Program (CEX2018-000806-S), and support from the Generalitat de Catalunya through the CERCA Program.

## Disclaimer

The findings and conclusions in this report are those of the authors and do not necessarily represent the views of the US Centers for Disease Control and Prevention.

