## Supplement for "Knowledge of COVID-19 Symptoms, Transmission, and Prevention: Evidence from Health and Demographic Surveillance in Southern Mozambique"

**Additional File 1**

**Supplemental methods**

*Wealth index generation*

A wealth index was generated based on a collection of assets and construction materials for the main dwelling of a given household. To generate the index, we followed recommendations from the DHS and the World Food Programme (WFP) that summarize steps for calculating an asset based wealth index [1-3], including coding instructions for Stata [4], and an adaptation of this coding process implemented in R [5, 6]. Using these documents to guide us, we generated our wealth index by identifying a list of household assets for inclusion in our index computation, recoded all household assets into dichotomous variables; recoded dwelling materials into improved vs. non-improved dichotomous variables; and assessed level of representation of a given variable within the population (per WFP recommendations, a given variable was included in further calculations if percent ownership ranged between 5 and 95 percent); employed principal components analysis (PCA) with varimax rotation to calculate component scores for those households living in either rural or urban areas, which explained 45% of the variation in both subsamples; extracted and combined the PCA scores of the first component from the urban and rural subsamples; and finally organized the scores into wealth quintiles to generate a composite asset index. The distribution of assets owned by households as well as the materials used for constructing a household’s residence are presented as percentages in Figure S1. The wealth quintiles for rural, urban, and combined are presented in Figure S2.

*Principal components factor analysis*

We used principal components factor analysis to create scores for assessing the degree of knowledge of COVID-19 symptoms based on nine variables, transmission based on nine variables, and prevention based on ten variables. The overall Kaiser-Meyer-Olkin index of sampling adequacy was 0.71, 0.73, and 0.83 for knowledge of symptoms, transmission, and prevention, so we concluded the sample size and data were adequate for the PCAs [16]. Knowledge of COVID-19 symptoms variables included: dry cough, headaches, fever, flu-like symptoms, difficulty breathing, sore throat, cough with sputum, muscle pain, and vomiting. Knowledge of COVID-19 transmission variables included: droplets from an infected person, inhalation of air from an infected person, touching an infected person’s hands, touching a fomite, touching an infected person, hugging an infected person, kissing an infected person, touching a fomite and then touching your mouth, touching a fomite and then touching your eyes or nose, touching your mouth, touching your nose, and touching your eyes. ‘Touching a fomite and then touching your mouth’ and ‘touching your mouth’ were collapsed to ‘touching your mouth,’ and ‘touching a fomite and then touching your eyes or nose,’ ‘touching your nose,’ and ‘touching your eyes’ were collapsed to ‘touching your eyes or nose’ as these variables were found to be correlated. Knowledge of COVID-19 prevention variables included: always wash hands with soap, always wear a mask, social distancing, wash hands with alcohol always, avoid crowded places, avoid traveling, avoid touching mouth, avoid touching nose, and avoid touching eyes. The assessment of knowledge started by assigning scores of one to an option if that option was mentioned by a particular respondent, and zero if he/she did not mention it. Individuals who did not know or did not respond to a knowledge question were assigned a score of zero for that question. The resultant compound factor for knowledge of symptoms included seven variables that accounted for 32% of the variability in the data: difficulty breathing, dry cough, fever, headaches, muscle pain, and sore throat (Table S1). Cough with sputum, flu-like symptoms, and vomiting had low correlation with the component and were dropped from the PCA to improve performance. The resultant compound factor for knowledge of transmission included all variables except ‘inhalation of air from an infected person’ that explained 29% of the data variability. The resultant compound factor for knowledge of prevention included eight variables that explained 31% of the data variability: avoid crowded places, touching eyes, touching mouth, touching nose, or traveling; social distancing; quarantine; and wash hands with alcohol. Washing hands with soap and wearing a mask were not included. Subsequent factors explained little variability. Therefore, only the first factor was retained from each PCA. These variables were then weighted against their eigenvector coefficients. Knowledge of symptoms, transmission, and prevention scores ranged from 0 to 3.3, 0 to 4.2, and 0 to 4.7, respectively, with higher scores representing greater knowledge. We used Pearson’s correlation to evaluate associations between the three knowledge indices.

*Multiple Imputation*

We used multiple imputation with predictive mean matching for these missing data to retain statistical power and avoid selection bias. In the imputation model, we included all individual-level (sex, age group, religion, education, occupation, marital status, had COVID-19 symptoms) and household-level indicators (household size, number of children under 5 years, number of adults over 60 years, number of pregnant women, wealth index), heard of coronavirus, sources of information, management of suspected cases, and knowledge of symptoms, transmission, and prevention indices. We created 20 imputed datasets, each using 20 cycles of regression switching, and used Rubin’s rules to combine the estimated regression coefficients and variances from the completed datasets [7].

*Poisson and logistic regression*

Quasi-Poisson regression was used to evaluate unadjusted and adjusted associations between characteristics (age; sex; language; religion; marital status; education; occupation; wealth index; had COVID-19 symptoms; number of household members, children under 5, older adults, and pregnant women; sources of information) and outcomes (knowledge of symptoms, transmission, and prevention scores) (Supplemental Methods). Additionally, logistic regression was used to evaluate unadjusted and adjusted associations between the same characteristics and households reporting that a household member had experienced symptoms associated with COVID-19. Variables were selected for inclusion in the final adjusted regression models if their unadjusted associations with outcomes (knowledge of symptoms, transmission, and prevention scores; had COVID-19 symptoms) had theoretical justification. Other studies have demonstrated significant associations between COVID-19 knowledge and age [8], sex [9], language, religion [10], marital status [11], education [12], occupation [10], income [10], had COVID-19 symptoms [13], household size [14], children in household [15], older adults [16], and sources of information [16]. Therefore, all variables from unadjusted analyses were included in the final model. Values of *P*<0.05 were considered statistically significant. We used the “car” package in R to derive generalized variance inflation factors (GVIF^(1/(2*df)^) for all independent variables in the adjusted model, where df is the degrees of freedom associated with each term [17].

*Mediation analysis*

We assessed whether sources of COVID-19 information mediated the relationship between educational attainment and knowledge of symptoms, transmission, and prevention scores. This analysis followed causal mediation analysis methods as previously described by VanderWeele [18]. The mediation models were binomial models to estimate the association between education and sources of information (TV, radio, Short Message Service [SMS]/WhatsApp, hospital, community leaders), which are dichotomous variables. We estimated quasi-Poisson models for the association between education and knowledge scores, adjusting for the mediators. The “mediation” package in R 4.2.3 statistical software (R Foundation for Statistical Computing) was used for multilevel causal mediation analyses [19]. We ran 1,000 Monte Carlo simulations in this analysis for variance estimation. Estimates, standard errors, and the proportion mediated are reported.

| Table S1. Principal components factor analysis of knowledge variables, Mozambique, September 2021 – January 2022 (N=33,087). | |
| --- | --- |
| Characteristic | Factor Pattern |
| Knowledge of symptoms |  |
| Difficulty breathing | 0.55 |
| Dry cough | 0.61 |
| Fever | 0.58 |
| Headaches | 0.61 |
| Muscle pain | 0.39 |
| Sore throat | 0.60 |
| *Eigenvalue* | 1.89 |
| Explained variance | 32% |
| Knowledge of transmission |  |
| Droplets from an infected person | 0.44 |
| Hugging an infected person | 0.63 |
| Kissing an infected person | 0.62 |
| Touching a fomite | 0.41 |
| Touching an infected person | 0.45 |
| Touching an infected person’s hands | 0.49 |
| Touching your eyes or nose | 0.59 |
| Touching your mouth | 0.63 |
| *Eigenvalue* | 2.32 |
| Explained variance | 29% |
| Knowledge of prevention |  |
| Avoid crowded places | 0.46 |
| Avoid touching eyes | 0.81 |
| Avoid touching mouth | 0.80 |
| Avoid touching nose | 0.81 |
| Avoid traveling | 0.60 |
| Quarantine | 0.53 |
| Social distancing | 0.33 |
| Wash hands with alcohol always | 0.40 |
| *Eigenvalue* | 3.09 |
| Explained variance | 39% |

| Table S2. Had symptoms suggestive of COVID-19 since COVID-19 was first reported in Mozambique (N=2,465) | | |
| --- | --- | --- |
| Symptom | N | % (95% CI) |
| Flu-like symptoms | 1242 | 50.4 (48.4, 52.4) |
| Dry cough | 1182 | 48.0 (46.0, 49.9) |
| Headaches | 832 | 33.8 (31.9, 35.7) |
| Fever | 665 | 27.0 (25.2, 28.8) |
| Cough with sputum | 541 | 21.9 (20.3, 23.6) |
| Sore throat | 194 | 7.9 (6.9, 9.0) |
| Muscle pain | 186 | 7.5 (6.5, 8.7) |
| Difficulty breathing | 162 | 6.6 (5.6, 7.6) |
| Vomiting | 18 | 0.7 (0.4, 1.2) |
| Other | 78 | 3.2 (2.5, 4.0) |
| CI: confidence interval | | |

| Table S3. Sources of information about COVID-19, Mozambique (N=33,087), | | |
| --- | --- | --- |
| Source | N | % (95% CI) |
| TV | 14551 | 44.0 (43.4, 44.5) |
| Community leaders | 11980 | 36.2 (35.7, 36.7) |
| Radio | 11134 | 33.7 (33.1, 34.2) |
| Hospital | 10518 | 31.8 (31.3, 32.3) |
| SMS/WhatsApp | 6748 | 20.4 (20.0, 20.8) |
| CI: confidence interval | | |

| **Table S4**. Mediation of sources of COVID-19 information on the association between educational attainment and knowledge of symptoms index^a^ derived from principal components analysis, Mozambique, September 2021 – January 2022 (N=33,087) | | | | | |
| --- | --- | --- | --- | --- | --- |
|  |  | Controlled direct effect | Natural indirect effect | Total effect |  |
| Education | Characteristic | Estimate (95% CI) | Estimate (95% CI) | Estimate (95% CI) | Proportion mediated |
| Higher | TV | 1.15 (1.02, 1.28) | 0.41 (0.38, 0.45) | 1.35 (1.22, 1.48) | 0.31 |
| Technical | TV | 1.03 (0.92, 1.17) | 0.36 (0.33, 0.39) | 1.21 (1.10, 1.34) | 0.30 |
| Secondary | TV | 0.66 (0.63, 0.70) | 0.22 (0.20, 0.23) | 0.79 (0.76, 0.82) | 0.27 |
| Primary | TV | 0.31 (0.28, 0.34) | 0.07 (0.06, 0.07) | 0.36 (0.34, 0.39) | 0.19 |
| Higher | Hospital | 1.35 (1.23, 1.49) | -0.01 (-0.02, -0.01) | 1.35 (1.22, 1.48) | -0.01 |
| Technical | Hospital | 1.22 (1.10, 1.35) | -0.01 (-0.02, -0.01) | 1.21 (1.10, 1.34) | -0.01 |
| Secondary | Hospital | 0.79 (0.76, 0.83) | -0.01 (-0.01, -0.01) | 0.79 (0.76, 0.82) | -0.01 |
| Primary | Hospital | 0.36 (0.34, 0.39) | -0.01 (-0.01, 0.00) | 0.36 (0.34, 0.39) | -0.01 |
| Higher | Radio | 1.32 (1.19, 1.45) | 0.07 (0.05, 0.10) | 1.35 (1.22, 1.48) | 0.05 |
| Technical | Radio | 1.19 (1.07, 1.32) | 0.06 (0.04, 0.09) | 1.21 (1.10, 1.34) | 0.05 |
| Secondary | Radio | 0.77 (0.74, 0.80) | 0.04 (0.03, 0.05) | 0.79 (0.76, 0.82) | 0.05 |
| Primary | Radio | 0.34 (0.31, 0.37) | 0.03 (0.03, 0.04) | 0.36 (0.34, 0.39) | 0.09 |
| Higher | SMS/WhatsApp | 1.22 (1.09, 1.35) | 0.29 (0.26, 0.34) | 1.35 (1.22, 1.48) | 0.22 |
| Technical | SMS/WhatsApp | 1.09 (0.97, 1.22) | 0.27 (0.23, 0.31) | 1.21 (1.10, 1.34) | 0.22 |
| Secondary | SMS/WhatsApp | 0.71 (0.68, 0.75) | 0.14 (0.13, 0.15) | 0.79 (0.76, 0.82) | 0.18 |
| Primary | SMS/WhatsApp | 0.34 (0.31, 0.37) | 0.03 (0.03, 0.04) | 0.36 (0.34, 0.39) | 0.09 |
| Higher | Community leaders | 1.38 (1.25, 1.50) | -0.07 (-0.09, -0.05) | 1.35 (1.22, 1.48) | -0.05 |
| Technical | Community leaders | 1.25 (1.14, 1.38) | -0.10 (-0.11, -0.08) | 1.21 (1.10, 1.34) | -0.08 |
| Secondary | Community leaders | 0.82 (0.79, 0.86) | -0.07 (-0.08, -0.06) | 0.79 (0.76, 0.82) | -0.09 |
| Primary | Community leaders | 0.38 (0.35, 0.41) | -0.02 (-0.03, -0.02) | 0.36 (0.34, 0.39) | -0.07 |

^a^ Knowledge of symptoms index included: difficulty breathing, dry cough, fever, headaches, muscle pain, and sore throat

| **Table S5**. Mediation of sources of COVID-19 information on the association between educational attainment and knowledge of transmission index^a^ derived from principal components analysis, Mozambique, September 2021 – January 2022 (N=33,087) | | | | | |
| --- | --- | --- | --- | --- | --- |
|  |  | Controlled direct effect | Natural indirect effect | Total effect |  |
| Education | Characteristic | Estimate (95% CI) | Estimate (95% CI) | Estimate (95% CI) | Proportion mediated |
| Higher | TV | 1.21 (1.07, 1.34) | 0.44 (0.40, 0.49) | 1.37 (1.23, 1.51) | 0.32 |
| Technical | TV | 0.97 (0.85, 1.10) | 0.35 (0.32, 0.39) | 1.12 (1.00, 1.25) | 0.32 |
| Secondary | TV | 0.62 (0.59, 0.66) | 0.21 (0.20, 0.22) | 0.73 (0.70, 0.76) | 0.29 |
| Primary | TV | 0.31 (0.29, 0.34) | 0.06 (0.06, 0.07) | 0.35 (0.33, 0.38) | 0.18 |
| Higher | Hospital | 1.35 (1.25, 1.52) | -0.05 (-0.08, -0.03) | 1.37 (1.23, 1.51) | -0.04 |
| Technical | Hospital | 1.13 (1.02, 1.27) | -0.05 (-0.07, -0.03) | 1.12 (1.00, 1.25) | -0.05 |
| Secondary | Hospital | 0.75 (0.72, 0.78) | -0.04 (-0.05, -0.03) | 0.73 (0.70, 0.76) | -0.05 |
| Primary | Hospital | 0.36 (0.33, 0.38) | -0.01 (-0.01, 0.00) | 0.35 (0.33, 0.38) | -0.02 |
| Higher | Radio | 1.34 (1.20, 1.48) | 0.11 (0.07, 0.15) | 1.37 (1.23, 1.51) | 0.08 |
| Technical | Radio | 1.09 (0.97, 1.22) | 0.09 (0.06, 0.12) | 1.12 (1.00, 1.25) | 0.08 |
| Secondary | Radio | 0.71 (0.68, 0.74) | 0.05 (0.04, 0.06) | 0.73 (0.70, 0.76) | 0.07 |
| Primary | Radio | 0.33 (0.30, 0.36) | 0.04 (0.04, 0.05) | 0.35 (0.33, 0.38) | 0.12 |
| Higher | SMS/WhatsApp | 1.21 (1.07, 1.35) | 0.43 (0.37, 0.49) | 1.37 (1.22, 1.51) | 0.31 |
| Technical | SMS/WhatsApp | 0.96 (0.85, 1.10) | 0.36 (0.32, 0.41) | 1.12 (1.00, 1.26) | 0.33 |
| Secondary | SMS/WhatsApp | 0.64 (0.60, 0.67) | 0.19 (0.18, 0.20) | 0.73 (0.70, 0.76) | 0.26 |
| Primary | SMS/WhatsApp | 0.33 (0.30, 0.38) | 0.05 (0.04, 0.05) | 0.35 (0.33, 0.38) | 0.13 |
| Higher | Community leaders | 1.39 (1.26, 1.53) | -0.10 (-0.12, -0.07) | 1.37 (1.23, 1.51) | -0.08 |
| Technical | Community leaders | 1.15 (1.04, 1.29) | -0.12 (-0.15, -0.10) | 1.12 (1.00, 1.25) | -0.11 |
| Secondary | Community leaders | 0.77 (0.74, 0.80) | -0.09 (-0.10, -0.08) | 0.73 (0.70, 0.76) | -0.12 |
| Primary | Community leaders | 0.37 (0.35, 0.40) | -0.03 (-0.03, -0.02) | 0.35 (0.33, 0.38) | -0.08 |

^a^ Knowledge of transmission index included: droplets from an infected person, hugging an infected person, kissing an infected person, touching a fomite, touching an infected person, touching an infected person’s hands, touching your eyes or nose, and touching your mouth

| **Table S6**. Mediation of sources of COVID-19 information on the association between educational attainment and knowledge of prevention index^a^ derived from principal components analysis, Mozambique, September 2021 – January 2022 (N=33,087) | | | | | |
| --- | --- | --- | --- | --- | --- |
|  |  | Controlled direct effect | Natural indirect effect | Total effect |  |
| Education | Characteristic | Estimate (95% CI) | Estimate (95% CI) | Estimate (95% CI) | Proportion mediated |
| Higher | TV | 0.85 (0.74, 0.97) | 0.37 (0.33, 0.41) | 0.97 (0.86, 1.11) | 0.38 |
| Technical | TV | 0.72 (0.60, 0.84) | 0.31 (0.27, 0.35) | 0.83 (0.72, 0.95) | 0.37 |
| Secondary | TV | 0.42 (0.39, 0.45) | 0.17 (0.16, 0.18) | 0.50 (0.48, 0.53) | 0.34 |
| Primary | TV | 0.17 (0.15, 0.19) | 0.05 (0.04, 0.05) | 0.20 (0.18, 0.22) | 0.23 |
| Higher | Hospital | 0.99 (0.88, 1.11) | -0.08 (-0.12, -0.05) | 0.97 (0.86, 1.11) | -0.08 |
| Technical | Hospital | 0.86 (0.75, 0.96) | -0.09 (-0.12, -0.05) | 0.83 (0.73, 0.94) | -0.10 |
| Secondary | Hospital | 0.52 (0.50, 0.55) | -0.06 (-0.07, -0.05) | 0.50 (0.48, 0.53) | -0.12 |
| Primary | Hospital | 0.21 (0.19, 0.22) | -0.01 (-0.01, 0.00) | 0.20 (0.18, 0.22) | -0.04 |
| Higher | Radio | 0.94 (0.83, 1.07) | 0.11 (0.07, 0.14) | 0.97 (0.86, 1.10) | 0.11 |
| Technical | Radio | 0.81 (0.70, 0.92) | 0.09 (0.06, 0.12) | 0.83 (0.72, 0.94) | 0.11 |
| Secondary | Radio | 0.48 (0.46, 0.51) | 0.05 (0.04, 0.06) | 0.50 (0.48, 0.53) | 0.10 |
| Primary | Radio | 0.18 (0.16, 0.20) | 0.04 (0.03, 0.04) | 0.20 (0.18, 0.22) | 0.18 |
| Higher | SMS/WhatsApp | 0.84 (0.73, 0.96) | 0.39 (0.35, 0.45) | 0.97 (0.86, 1.11) | 0.40 |
| Technical | SMS/WhatsApp | 0.70 (0.60, 0.82) | 0.34 (0.29, 0.38) | 0.83 (0.72, 0.94) | 0.41 |
| Secondary | SMS/WhatsApp | 0.42 (0.39, 0.45) | 0.17 (0.16, 0.18) | 0.50 (0.47, 0.53) | 0.33 |
| Primary | SMS/WhatsApp | 0.18 (0.16, 0.20) | 0.04 (0.03, 0.04) | 0.20 (0.18, 0.22) | 0.18 |
| Higher | Community leaders | 1.01 (0.89, 1.13) | -0.15 (-0.19, -0.12) | 0.97 (0.86, 1.10) | -0.15 |
| Technical | Community leaders | 0.88 (0.77, 0.99) | -0.20 (-0.23, -0.16) | 0.83 (0.73, 0.95) | -0.24 |
| Secondary | Community leaders | 0.54 (0.52, 0.57) | -0.13 (-0.14, -0.12) | 0.50 (0.48, 0.53) | -0.26 |
| Primary | Community leaders | 0.22 (0.21, 0.24) | -0.04 (-0.04, -0.03) | 0.20 (0.19, 0.22) | -0.18 |

^a^ Knowledge of prevention index included: avoid crowded places, touching eyes, touching mouth, touching nose, or traveling; social distancing; quarantine; and wash hands with alcohol

| Table S7. Generalized variance inflation factors of independent variables for analysis of associations between demographic characteristics and knowledge of COVID-19, Mozambique, September 2021 – January 2022 (N=33,087). | |
| --- | --- |
| Variable | **Generalized variance inflation factor** |
| Education | 1.097399 |
| Age group | 1.170266 |
| Occupation | 1.046221 |
| Sex | 1.123760 |
| Religion | 1.024557 |
| Language | 1.012741 |
| Had COVID-19 symptoms | 1.021757 |
| Marital status | 1.106240 |
| Household size | 1.057320 |
| Adults over 60 | 1.158546 |
| Children under 5 | 1.117738 |
| Pregnant women | 1.008662 |
| Wealth index | 1.072566 |
| Community leaders | 1.121468 |
| Hospital | 1.099873 |
| Radio | 1.040220 |
| SMS/WhatsApp | 1.103482 |
| TV | 1.292671 |

**Figure S1.** Households’ Asset Ownership and Dwelling Construction Materials included in Wealth Index. A Mitad is a portable, electric hotplate used for cooking. Improved classifications for dwelling materials were based on DHS recommendations.

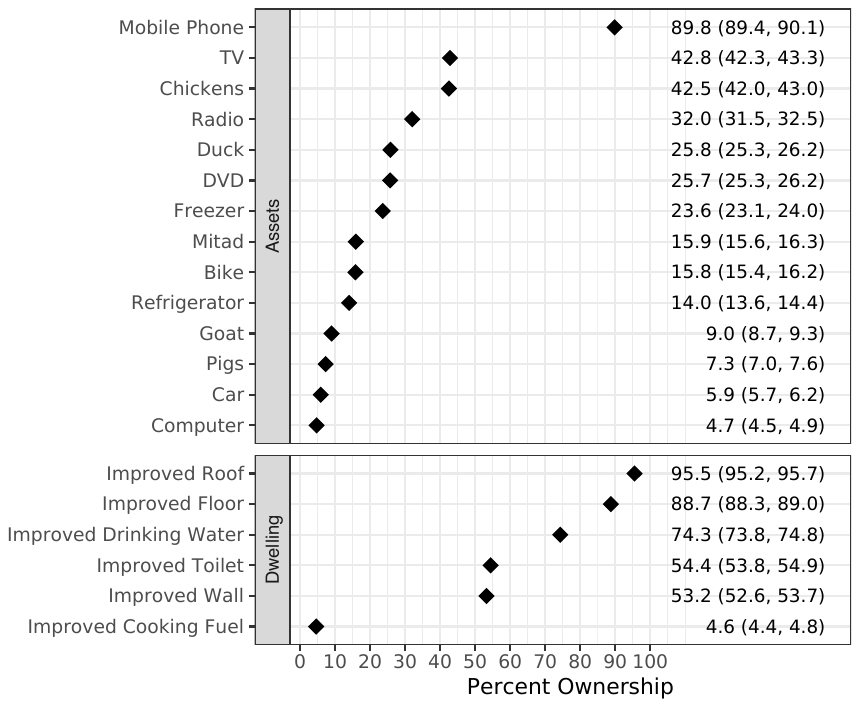

**Figure S2.** Household Wealth Index Quintile Distribution

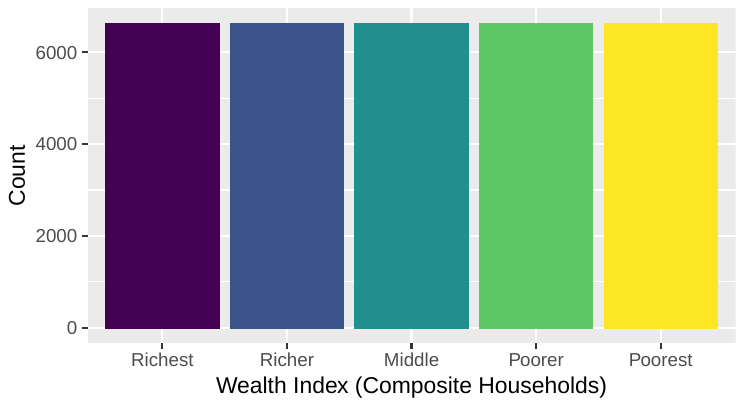

**Figure S3**. Unadjusted associations between demographics and sources of information, and knowledge of prevention, symptoms, and transmission indices^a^ derived from principal components analysis, Mozambique, September 2021 – January 2022 (N=33,087).

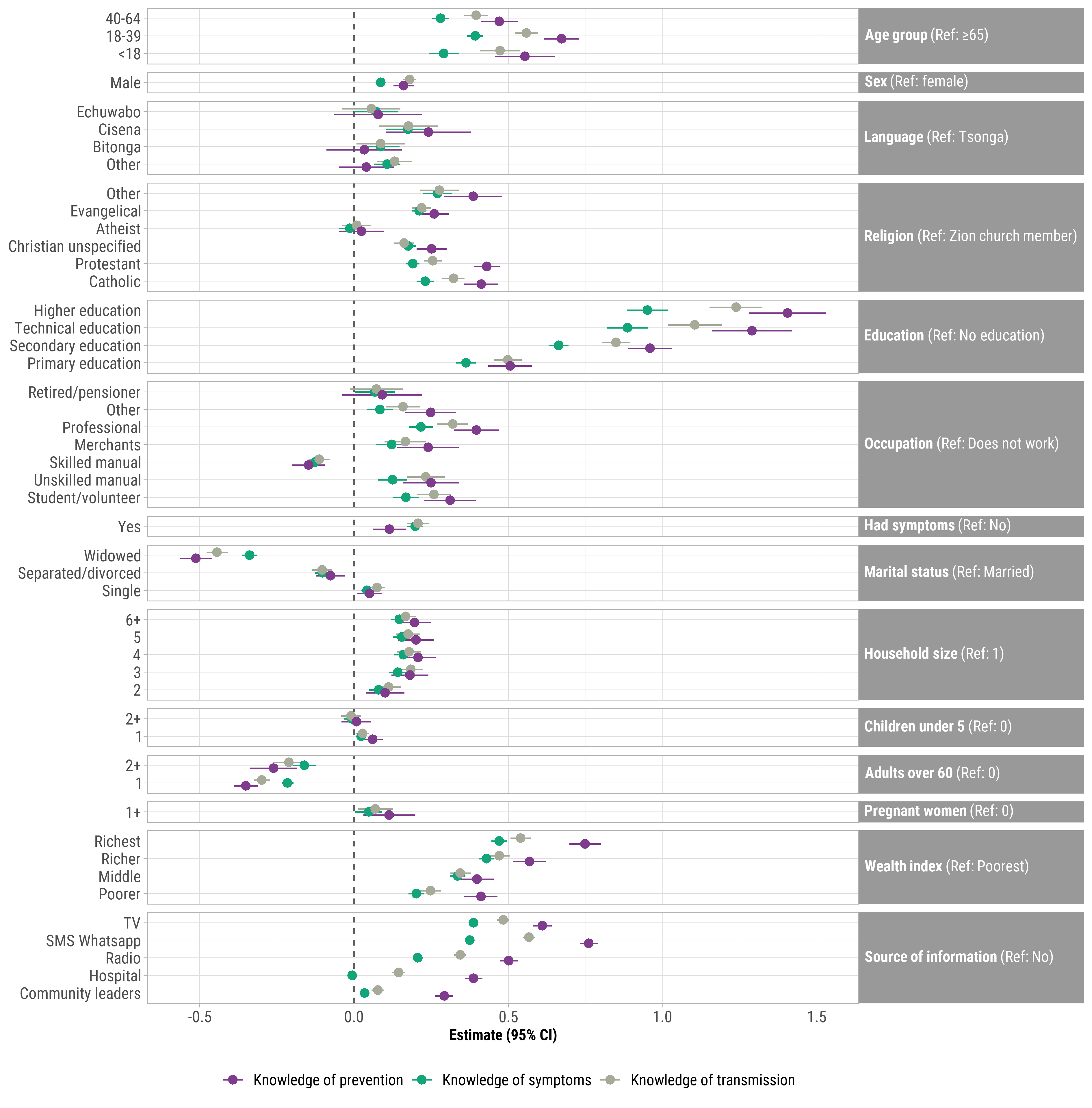
Points represent β coefficients and error bars represent 95% confidence intervals.
^a^ Knowledge of prevention includes: avoid crowded places, touching eyes, touching mouth, touching nose, or traveling; social distancing; quarantine; and wash hands with alcohol. Knowledge of symptoms includes: difficulty breathing, dry cough, fever, headaches, muscle pain, and sore throat. Knowledge of transmission includes: avoid crowded places, touching eyes, touching mouth, touching nose, or traveling; social distancing; quarantine; and wash hands with alcohol.

**Figure S4**. Pearson’s correlation between principal components analysis-derived knowledge of COVID-19 symptoms, transmission, and prevention scores. Coefficients (*r*) and 95% confidence intervals are shown.

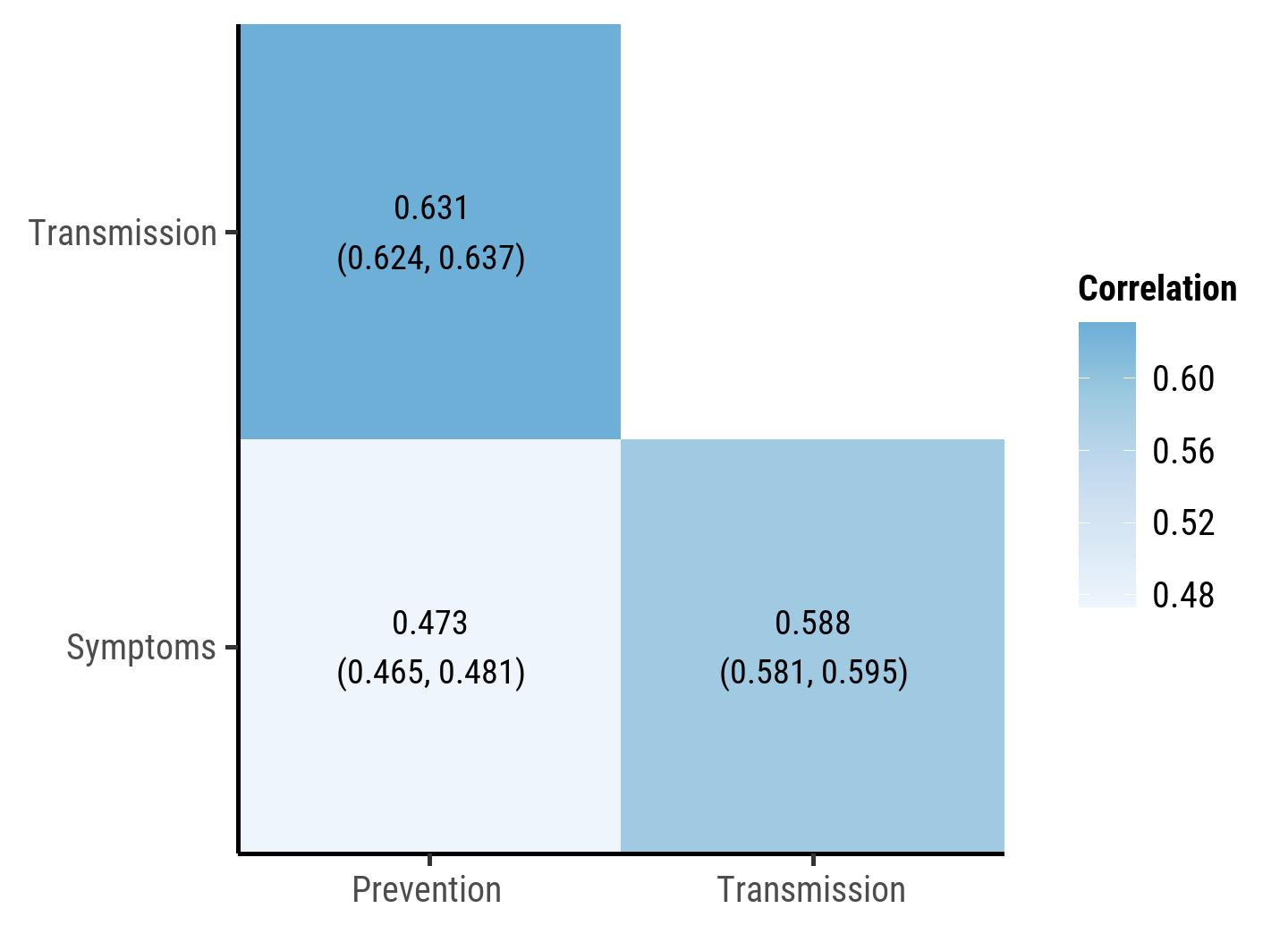
